# Knowledge, attitudes and practices towards antibiotic use and resistance in Kyegegwa district, Uganda – a questionnaire study

**DOI:** 10.1101/2023.04.06.23288253

**Authors:** Mary Ann Kahunde, Terence Odoch, David Okello Owiny, Clovice Kankya, Marisa B. Kaelin, Sonja Hartnack

## Abstract

**Background:** Antimicrobials are widely used to protect human and animal health. Wide scale misuse can lead to the development and spread of antimicrobial resistance (AMR). In low and middle-income countries, knowledge, attitudes and practices are assumed to contribute to AMR.

**Objective:** To provide empirical data on knowledge and attitudes towards antibiotic use and resistance in livestock farmers, human and animal health professionals in Kyegegwa district, Uganda. To assess which factors (farming, education, affiliation with health-related ministries, gender, keeping cattle, goats, pigs, poultry or sheep) are associated with poor knowledge.

**Methods:** A questionnaire related to antibiotic usage (AMU) and AMR, as well as demographic data was developed and administered to health practitioners of both the humanmedical and the livestock sector, and (semi)-intensive livestock farmers. Polytomous latent class analysis was used to cluster respondents - based on their responses - in different classes of knowledge and attitudes. The association between the probability of belonging to latent classes and demographic data was assessed by multinomial regression models and conditional inference trees.

**Results:** In total 1022 responses were available (response rate 68.1%). More than 50% of the cattle received antibiotics at least three times during the last six months and in 39.1% members of the respondents’ households of this study were on antibiotics. Three latent classes related to attitude towards antibiotic access, usage and disposal; and two classes regarding resistance have been revealed. Class membership was associated with a number of socio-demographic factors.

**Conclusion:** Inadequate knowledge and inappropriate practices as evidenced in this study should support further policy formulations and strategies to regulate AMU. It will also be useful in refining the implementation of local and national action plans and efforts to control AMR. A key component of this will require promotion of One Health approach and effective communication to tackle prevailing misconceptions.

## Introduction

Antimicrobials are widely used to protect human and animal health from pathogens. Antimicrobials are pre-requisites for modern healthcare as first line of treatment for many infectious diseases, and for the management of secondary infections during surgical operations. Unfortunately, wide scale misuse and abuse has led to development of antimicrobial resistance (AMR) in microorganisms (1–4). It has been estimated that 20 – 50% of antibiotics used is either unnecessary or inappropriate (5,6). Consequently, AMR has become a global public health threat. It is estimated, that by 2050 approximately 10 million people will die annually due to infections related to antibiotic resistance (7). In addition, AMR is likely to significantly have negative effects on the global economy - mainly in low and middle-income countries (LMICs) (5). While development of AMR is a natural process that has been long recognized, its spread is exacerbated by many factors, some of them related to inappropriate prescription and use (8–10).

In LMICs, one of the factors contributing to misuse of antimicrobials is related to knowledge attitudes and practices of the key players in antimicrobial usage. Many studies have reported that antibiotic regime non-adherence and inappropriate antibiotic use are strongly associated with public awareness and knowledge of antibiotics (11–14). Unfortunately, limited studies have examined the knowledge attitudes and practices of users and prescribers of antimicrobials in developing countries (15). The main aim of this study was to provide empirical data on knowledge and attitudes towards antibiotic use and resistance of livestock farmers, human and animal health professionals in Kyegegwa district, Uganda.

## Material and methods

### Questionnaire design

A questionnaire was developed based on the questions from Vallin (16) aiming to investigate knowledge and attitudes regarding antibiotic use and resistance in Sweden. To adapt the questions to the local situation in Uganda, the content was presented and discussed in a participatory approach with participants of medical, veterinary and pharmaceutical background during the 2017 Dialogue Days on Global Health Challenges between Makerere University and University of Zurich.

The questionnaire consisted of four parts: (i) antibiotic consumption (8 questions on animals and 9 on humans), (ii) antibiotic use (12 questions), (iii) antibiotic misconceptions (17 questions), and (iv) side effects and resistance (7 questions). The possible answers consisted of (“yes”, “no”, “don’t know”), multiple options to tick and related to statements (“I agree”, “I do not agree”, “I do not know”). Demographic information included questions on gender, age, education, livestock enterprise, and enterprise type. The questionnaire was developed in English and translated into the local language Rutooro upon administration. It was analysed and revised by three independent supervisors and pre-tested on members of the target population who were not repeated on the final investigation. The questionnaire is available in the supplementary material (S1 Questionnaire).

### Study area and population

Kyegegwa District in Uganda is bordered by Kibale District to the north, Mubende District to the east, Kiruhura District to the south, Kamwenge District to the south-west, Sembabule to the south-east and Kyenjojo District to the north-west. It is divided into 14 lower local governments (LLGs) with nine rural Sub-Counties, five town councils, 48 parishes, 19 town wards and 266 villages. It is located approximately 110 kilometres (68 miles), east of Fort Portal, Kabarole District, and is part of Tooro Sub region. Kyegegwa District was created by an act of the Uganda Parliament on 1 July 2009. Before then, it was part of Kyenjojo District, which is also part of the Tooro Sub region, from Tooro Kingdom.

According to the Uganda Bureau of Statistics (17), the projected population of Kyegegwa district was 417’000 people. The economic activities in the district include livestock farming, cultivation (maize, bananas, fruits, beans, groundnuts, cassava, millet, Irish potatoes and sweet potatoes), aquaculture or fish farming, business in agricultural produce and apiary.

### Data collection

The study targeted 1500 participants from 6 Sub-counties of Ruyonza, Rwentuha, Mpara, Kyegegwa T/C, Kakabara and Kyegegwa Sub-counties of Kyegegwa District in autumn 2018. Included were public and private health practitioners of both the medical and the livestock sector, with intensive and semi-intensive livestock farmers, aged 18-75 years.

Percentages were purposively used for the inclusion of individual participants within the 6 Sub-counties (based on those areas laying most in the cattle corridor and those not in the cattle corridor). The 1500 participants were selected based on the population size of the study category and level of antibiotic use in regard to exposure to antibiotic resistance (Personal field experience). The aim was to select 50% livestock farmers (25% intensive and 25% semi-intensive), 25% human public and private health practitioners, 25% veterinary public and private practitioners.

Interviews were self-administered by selected Sub-county extension staff who were pre-trained and guided by the researcher (MAK) before data collection. Health workers were organized in common places for every health unit, and questionnaires administered at once after guidance and so were the village health team workers at every parish level, from which sensitization and training on antibiotic resistance were carried out. Farmers were organised in community meetings, sensitized and questionnaires self-administered with assistance from the trained veterinarians. Farm visits and home to home interviews were made with random sampling for conclusive and evident data collection.

### Data management and analysis

The data were entered manually in an excel sheet. Data cleaning and the statistical analysis was performed with the open-source software R version 4.1.3 (18). For the description of the demographic variables and the questions related to knowledge and attitudes towards antibiotic usage and resistance, multinomial and Jeffreys binomial 95% confidence intervals were obtained with the commands MultinomCI() and BinomCI() in the package DescTools (19). With the aim to detect latent classes in the responses related to the attitude towards antibiotic access, usage and disposal as well as towards resistance, polytomous latent class analyses were performed with the command poLCA() in the poLCA package (20). Subsequently, 1 to 5 classes were assessed with 20 repetitions and 50 000 iterations. Selection of the final model was based on BIC and having at least 10% of the respondents in each class. The resulting latent classes were considered as outcome variable in multinomial regression analyses. The following nine factor variables were included both in a univariable and a multivariable approach: livestock farming (none as baseline, intensive and semi-intensive), affiliation with a ministry (no as baseline, MAAIF. Ministry of Agriculture, Animal Industry and Fisheries; MoH, Ministry of Health), gender (male as baseline, and female), education (primary as baseline, ordinary, tertiary and university), keeping cattle, goats, pigs, poultry and sheep (baseline always no). With the aim to predict which of the nine factor variables is most closely related to the predicted latent class membership, conditional inference trees, relying on a recursive partitioning algorithm were performed with the command ctree() from the package partykit (21).

### Ethics

Clearance for the study was sought from the School of Biosecurity, Biotechnology and Laboratory Sciences (SBLS) Institutional Review Board, and the College of Veterinary Medicine, Animal Resources and Biosecurity (COVAB), Makerere University. Permission to conduct this study was sought from the Chief Administrative Officer and the District Council of Kyegegwa District Local Government. Verbal consent was sought from the respondents who were medical and veterinary professionals and livestock farmers, and confidentiality of the information was guaranteed before the interview.

## Results

### Response rate

In total 1022 responses were available with 319 (31.3%) respondents being affiliated with the MOH, 41 (4%) with the MAAIF and 660 (64.7%) being not affiliated with a ministry (2 NA). The response rate was 68.1 %. The MOH employees comprised 165 (51.7%) VHT (village health team), 118 (37.0%) nurses, 15 (4.7%) public health officer, 12 (3.8%) clinical officer, five (1.6%) doctors and four (1.2%) pharmacists. The MAAIF employees comprised 38 (92.7%) animal husbandry assistants, two (4.9%) animal health officers and one (2.4%) veterinarian.

Out of MOH employees, 27 (8.4%) described themselves as doing intensive as well as semi-intensive livestock farming and 265 (83%) indicated no livestock farming activity. Amongst MAAIF employees, six (14.6%) and 14 (34.1%) identified themselves as intensive and semi-intensive livestock farmer, while 21 (51.2%) did not act as livestock farmer. Amongst the respondents not being associated with a ministry, 216 (32.7%) were intensive and 444 (67.3 %) semi-intensive livestock farmers. Demographic data of the respondents including education, gender, as well as keeping cattle, goats, pigs, poultry and sheep are presented in Table 1.

**Table 1.** Demographic data of respondents classified for affiliation with a ministry and with type of farming

|  | Ministr<br>y |  |  | Farmer<br>(1 NA*) |  |  | Total |
| --- | --- | --- | --- | --- | --- | --- | --- |
|  | (2 NA*) |  |  |  |  |  |  |
| **Characteristic** | MAAIF<br>n (%)<br>[95%CI] | MOH<br>n (%)<br>[95%CI] | No<br>n (%)<br>[95%CI] | I<br>n (%)<br>[95%CI] | SI<br>n (%)<br>[95%CI] | No<br>n (%)<br>[95%CI] |  |
| **Education**<br>(7 NA*) |  |  |  |  |  |  |  |
| Primary | 8 (19.5)<br>[7.3;35.3]<br>] | 75<br>(23.6)<br>[17.9;29.6]<br>6] | 350<br>(53.5)<br>[49.5;57.5]<br>5] | 139<br>(56.8)<br>[50.2;62.8]<br>] | 243<br>(50.4)<br>[45.8;55.2]<br>2] | 51<br>(17.9)<br>[11.9;24.1]<br>1] | 433<br>(42.7)<br>[39.4;46.0]<br>0] |
| Ordinary | 1 (2.4)<br>[0;18.2]<br>] | 97(30.5)<br>[24.8;36.6]<br>6] | 203<br>(31.0)<br>[27.1;35.1]<br>1] | 58<br>(23.5)<br>[17.0;30.0]<br>0] | 162<br>(33.6)<br>[29.0;38.4]<br>4] | 81<br>(28.4)<br>[22.4;34.7]<br>7] | 302<br>(29.7)<br>[26.5;33.1]<br>1] |
| Tertiary | 26<br>(63.4)<br>[51.2;79.2]<br>2] | 120<br>(37.7)<br>[32.0;43.8]<br>8] | 71<br>(10.8)<br>[6.9;14.8]<br>] | 39<br>(15.8)<br>[9.7;22.3]<br>] | 51<br>(10.6)<br>[6.0;15.4]<br>] | 128<br>(44.9)<br>[38.9;51.1]<br>1] | 218<br>(21.5)<br>[18.2;24.9]<br>9] |
| University | 6 (14.6)<br>[2.4;30.4]<br>] | 26 (8.2)<br>[2.5;14.3]<br>] | 30 (4.6)<br>[0.6;8.6]<br>] | 11 (4.4)<br>[0;11]<br>] | 26 (5.4)<br>[5.4;10.2]<br>] | 25<br>(28.4)<br>[22.4;34.7]<br>7] | 62 (6.1)<br>[2.8;9.5]<br>] |
| **Gender**<br>(1 NA*) |  |  |  |  |  |  |  |
| Female | 9 (21.9)<br>[12.2;35.5] | 163<br>(51.1)<br>[45.4;56.7] | 219<br>(33.3)<br>[29.6;36.9] | 82<br>(33.1)<br>[27.4;39.2] | 163<br>(33.5)<br>[29.4;38.0] | 147<br>(51.4)<br>[45.4;57.3] | 393<br>(38.7)<br>[35.8;41.7] |
| Male | 32<br>(78.0)<br>[68.3;91.6] | 156<br>(48.9)<br>[43.2;54.4] | 440<br>(66.8)<br>[63.1;70.5] | 166<br>(66.9)<br>[61.3;73.1] | 323<br>(66.5)<br>[62.3;70.9] | 139<br>(48.6)<br>[42.6;54.6] | 628<br>(61.9)<br>[0.59;64.8] |
| **Cattle**<br>(16 NA*) |  |  |  |  |  |  |  |
| Yes | 30<br>(76.9)<br>[66.7;91.1] | 78<br>(25.5)<br>[20.9;30.6] | 434<br>(65.7)<br>[62.1;69.5] | 165<br>(66.5)<br>[60.9;72.7] | 317<br>(65.2)<br>[60.9;69.5] | 60<br>(22.0)<br>[17.3;26.9] | 542<br>(0.54)<br>[0.51;56.9] |
| No | 9 (23.2)<br>[12.8;37.3] | 228<br>(74.5)<br>[69.9;79.6] | 226<br>(34.2)<br>[30.6;38.0] | 83<br>(33.5)<br>[27.8;39.7] | 169<br>(34.8)<br>[30.4;39.1] | 212<br>(77.9)<br>[73.2;82.8] |  |
| **Goats**<br>(16 NA*) |  |  |  |  |  |  |  |
| Yes | 23<br>(59.0)<br>[46.1;76.1] | 93<br>(30.4)<br>[25.3;38.5] | 408<br>(61.8)<br>[58.0;65.7] | 151<br>(60.9)<br>[54.8;67.2] | 308<br>(63.3)<br>[59.0;67.8] | 65<br>(23.9)<br>[19.1;29.1] | 524<br>(52.1)<br>[49.0;55.2] |
| No | 16<br>(41.0)<br>[28.2;58.1] | 213<br>(69.6)<br>[64.7;75.1] | 252<br>(38.2)<br>[34.4;42.0] | 97<br>(39.1)<br>[33.1;45.3] | 178<br>(36.6)<br>[32.2;41.1] | 207<br>(76.1)<br>[71.3;81.3] |  |
| **Pigs**<br>(16 NA*) |  |  |  |  |  |  |  |
| Yes | 14<br>(35.9)<br>[23.1;52.1] | 67<br>(21.9)<br>[17.6;26.7] | 329<br>(49.8)<br>[45.9;53.8] | 86<br>(34.7)<br>[29.0;41.0] | 271<br>(55.8)<br>[51.2;60.4] | 54<br>(19.8)<br>[15.4;24.7] | 411<br>(40.8)<br>[37.8;43.9] |
| No | 25<br>(64.1)<br>[51.3;80.3] | 239<br>(78.1)<br>[73.8;82.9] | 331<br>(50.1)<br>[46.2;54.1] | 162<br>(65.3)<br>[59.7;71.6] | 215<br>(44.2)<br>[39.7;48.8] | 218<br>(80.1)<br>[75.7;85.0] |  |
| **Poultry**<br>(16 NA*) |  |  |  |  |  |  |  |
| Yes | 22<br>(56.4)<br>[43.6;73.8] | 84<br>(27.4)<br>[22.5;32.5] | 467<br>(70.7)<br>[67.3;74.3] | 178<br>(71.8)<br>[66.5;77.7] | 343<br>(70.6)<br>[66.7;74.8] | 53<br>(19.5)<br>[15.1;24.2] | 574<br>(53.9)<br>[50.9;56.9] |
| No | 17<br>(43.6)<br>[30.8;61.0] | 222<br>(72.4)<br>[67.6;77.6] | 193<br>(29.2)<br>[25.7;32.8] | 70<br>(28.2)<br>[23.0;34.1] | 143<br>(29.4)<br>[25.5;33.7] | 219<br>(80.5)<br>[76.1;85.3] |  |
| **Sheep** |  |  |  |  |  |  |  |
| (16 NA*) |  |  |  |  |  |  |  |
| Yes | 15<br>(38.5)<br>[25.6;<br>55.2] | 19 (6.2)<br>[3.9;8.9] | 173<br>(26.2)<br>[22.9;29.<br>6] | 62<br>(0.25)<br>[19.7;30.<br>4] | 128<br>(26.3)<br>[22.4;30.<br>3] | 17 (6.2)<br>[3.7;8.9] | 207<br>(20.6)<br>[18.2;23.<br>2] |
| No | 24<br>(61.5)<br>[48.7;78.<br>2] | 287<br>(93.8)<br>[91.5;96.<br>4] | 487<br>(73.8)<br>[70.4;77,<br>2] | 186<br>(75.0)<br>[69.7;80.<br>4] | 358<br>(73.7)<br>[69.7;77.<br>6] | 255<br>(93.7)<br>[91.2;96.<br>4] |  |
| **Total** | 41 (4.0)<br>[1.1;7.1] | 319<br>(31.3)<br>[28.3;34.<br>3] | 660<br>(64.7)<br>[61.8;67.<br>8] | 249<br>(28.0)<br>[24.8;31.<br>4] | 486<br>(47.6)<br>[44.3;51.<br>0] | 286<br>(24.4)<br>[21.1;27.<br>8] |  |

\*NA missing values

Asked about which antibiotics the respondent has ever used him- or herself, the following four antibiotics were mentioned most often: amoxicillin, followed by penicillin, erythromycin and co-trimoxazole. In animals, penicillin, tetracycline and gentamycin were mentioned most often (Table 2a). Amongst the 844 respondents which kept one of the following five species (cattle, goats, pigs, poultry or sheep), 147 kept one species, 247 kept two species, 231 kept three species, 171 kept four species and 48 kept all five species.

**Table 2a.** Antibiotics mentioned by the respondents to be used in humans and animals

| Antibiotic | Humans<br>n = 1022 |  |  | Animals<br>n = 844 |  |  |
| --- | --- | --- | --- | --- | --- | --- |
|  | n (%) | [95% CI] | NA | n (%) | [95% CI] | NA |
| Amoxicillin | 678 (66.5) | [63.6;69.4] | 3 | 107 (12.7) | [10.6;15.1] | 2 |
| Azithromycin | 212 (20.8) | [18.4;23.4] | 3 | 27 (3.2) | [2.2;4.6] | 2 |
| Cephalexin | 183 (17.9) | [15.7;20.4] | 2 | 36 (4.3) | [3.1;5.8] | 2 |
| Co-trimoxazole | 425 (41.7) | [38.7;44.7] | 3 | 100 (11.9) | [9.6;14.2] | 2 |
| Doxycyclin | 253 (24.8) | [22.2;27.5] | 3 | 221 (26.2) | [23.4;29.3] | 2 |
| Erythromycin | 429 (42.1) | [39.1;45.1] | 3 | 51 (6.1) | [4.6;7.8] | 2 |
| Gentamycin | 316 (31.0) | [28.2;33.9] | 3 | 303 (36.0) | [32.8;39.3] | 2 |
| Penicillin | 481 (47.2) | [44.1;50.3] | 3 | 585 (69.6) | [66.3;72.5] | 2 |
| Tetracycline | 334 (32.8) | [30.0;35.7] | 4 | 573 (68.1) | [64.8;71.1] | 2 |
| Trimethoprim | 160 (15.7) | [13.6;18.0] | 4 | 88 (10.4) | [8.5;12.6] | 2 |

In Table 2b the frequencies of antibiotic usage during the last six months in cattle, goats, pigs, poultry as well as different age categories of humans are presented. More than 50% of the cattle received antibiotics at least three times during the last six months, whereas goats, pigs, poultry and sheep received considerably less often antibiotics. In humans, the majority in all age categories received less than three times an antibiotic in the same time period.

**Table 2b.** Frequencies of antibiotic usage in different animal species and human age categories

| Species | Never | 1-2 times | 3-5 times | >5 times |
| --- | --- | --- | --- | --- |
|  | n (%)<br>[95% CI] | n (%)<br>[95% CI] | n (%)<br>[95% CI] | n (%)<br>[95% CI] |
| Cattle (15 NA*) | 27 (5.1)<br>[0.5;9.8] | 196 (37.2)<br>[32.6;41.9] | 186 (35.3)<br>[30.7;40.0] | 118 (22.4)<br>[17.8;27.1] |
| Goats (24 NA*) | 213 (42.6)<br>[38.0;47.3] | 190 (38.0)<br>[33.3;42.7] | 62 (12.4)<br>[7.8;17.1] | 35 (7.0)<br>[2.4;11.7] |
| Pigs (17 NA*) | 141 (35.8)<br>[30.7;41.2] | 164 (41.6)<br>[36.5;47.0] | 51 (12.9)<br>[7.9;18.3] | 38 (9.6)<br>[4.6;15.0] |
| Poultry (9 NA*) | 311 (55.0)<br>[50.9;59.5] | 175 (30.9)<br>[26.9;35.4] | 53 (9.4)<br>[3.1;13.8] | 26 (4.6)<br>[0.5;9.0] |
| Sheep (8 NA*) | 142 (71.3) | 29 (14.6) | 18 (9.0) | 10 (5.0) |
|  | [65.8;77.8] | [9.0;21.0] | [3.5;15.5] | [0;11.5] |
| Humans:<br>Respondent (31<br>NA*) | 333 (33.6)<br>[30.4;37.0] | 403 (40.7)<br>[37.4;44.1] | 118 (11.9)<br>[8.7;15.3] | 137 (13.8)<br>[10.6;17.2] |
| Humans: another<br>adult in same<br>household (88 NA*) | 389 (41.6)<br>[38.2;45.2] | 330 (35.3)<br>[32.0;38.9] | 109 (11.7)<br>[8.3;15.2] | 106 (11.3)<br>[8.0;14.9] |
| Children ≤ 2 years | 167 (25.4)<br>[21.5;29.5] | 315 (47.9)<br>[44.0;52.0] | 111 (16.9)<br>[12.9;20.9] | 64 (9.7)<br>[5.8;13.8] |
| Children 3-5 years | 255 (41.1)<br>[36.9;45.4] | 213 (34.3)<br>[30.2;38.6] | 112 (18.1)<br>[13.9;22.3] | 40 (6.4)<br>[2.2;10.7] |
| Children 6-12 years | 340 (53.0)<br>[49.1;57.2] | 204 (31.8)<br>[27.9;36.0] | 57 (8.9)<br>[5.0;13.0] | 40 (6.2)<br>[2.3;10.4] |
| Children 13-18 years | 325 (58.9)<br>[54.7;63.0] | 142 (25.7)<br>[21.5;29.9] | 49 (8.9)<br>[4.7;13.0] | 36 (6.5)<br>[2.3;10.7] |

\*NA missing values

Asked about the reasons to administer antibiotics to livestock in decreasing order the following answers were chosen: “when they are sick”, “to prevent disease”, “for growth and fattening”, and “when I feel it is necessary” (Table 2c). In decreasing order, the source of the antibiotic was the drug shop, veterinary doctor, market, pharmacy, neighbour, and mobile van. Most often the antibiotic was administered by the respondent or a family member, less often by animal health assistants, veterinary doctors, neighbours or livestock production officers. The question, whether a prescription for the antibiotic treatment of the animals was available was answered with “yes” by 335 respondents (43.7%, 95% CI [40.2;47.3]) and with “no” by 431 respondents (56.3%, 95% CI [52.7;59.7]). Observing a withholding period as recommended was answered with “yes” by 116 respondents (15.1%, 95% CI [12.7;17.8]), with “no” by 635 (82.9%, 95% CI [80.4;85.6]) and with “don’t know” by 15 (1.9%, 95% CI [0;4.7]).

**Table 2c.** Administration and source of antibiotics in livestock

| Question | “yes”<br>n (%)<br>[95% CI] | NA* |
| --- | --- | --- |
| When do you administer antibiotics to your livestock? |  |  |
| When they are sick | 692 (90.8) | 82 |
|  | [88.6;92.7] |  |
| To prevent disease | 485 (63.2)<br>[59.8;66.6] | 77 |
| For growth and fattening | 247 (32.1)<br>[28.9;35.5] | 75 |
| When I feel it is necessary | 166 (21.7)<br>[18.9;24.7] | 78 |
| What was the source of your antibiotic? |  |  |
| Drug shop | 508 (66.0)<br>[62.6;69.2] | 74 |
| Veterinary doctor | 221 (28.7)<br>[25.6;32.0] | 75 |
| Market | 172 (22.4)<br>[19.5;25.4] | 76 |
| Pharmacy | 100 (13.0)<br>[10.8;15.5] | 75 |
| Neighbour | 64 (8.3)<br>[6.5;10.4] | 76 |
| Mobile van | 21 (2.7)<br>[1.7;4.1] | 75 |
| Who administered for you the antibiotic? |  |  |
| Family member or myself | 555 (72.3)<br>[69.1;75.4] | 77 |
| Animal health assistant | 251 (32.7)<br>[29.4;36.0] | 76 |
| Veterinary doctor | 99 (12.9)<br>[10.7;15.4] | 79 |
| Neighbour | 72 (9.4)<br>[7.5;11.6] | 78 |
| Livestock production officer | 70 (9.2)<br>[7.3;11.4] | 80 |

\*NA missing values

In humans, whether anyone is currently taking antibiotics in the household, answered 378 (39.1%, 95% CI [36.0;42.4]) respondents with “yes”, 580 (60.0, 95% CI [56.9;63.3]) with “no” and 9 (0.9%, 95% CI [0;4.2]) with “don’t know” (NA 55).

Regarding the prescription, source and administration of antibiotics in humans, further details are presented in Table 2d. Information related to patient experience, patient-doctor relationship and antibiotic usage is presented in table 2e. Solely between 60 and 75% of the respondents have experienced antibiotic prescriptions for a family member or their animals by a doctor and even less by a pharmacy. A clear majority of the respondents agree that doctors provide adequate information and conduct thorough examinations if antibiotics are needed or not. Confidence in a doctor’s decision not to prescribe antibiotics was expressed by more than 70% of the respondents. While farmers and respondents affiliated with a ministry agreed in a similar way with most of the statements, for the statement that “a doctor who does not prescribe antibiotics when the patient thinks that they are needed, is not a good doctor” - a majority of the farmers agreed, but respondents affiliated with MAAIF/MOH did not.

**Table 2d.**
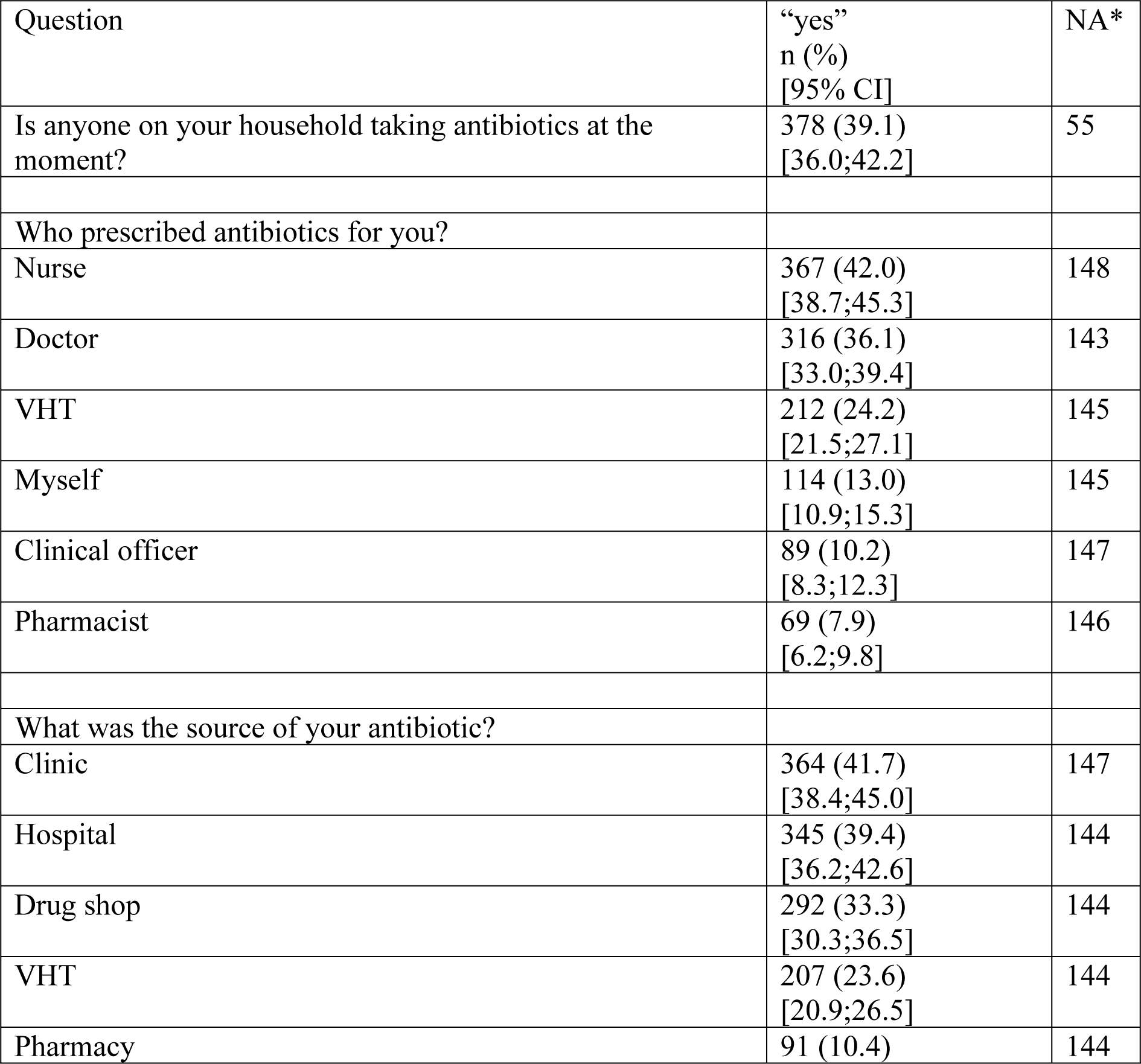
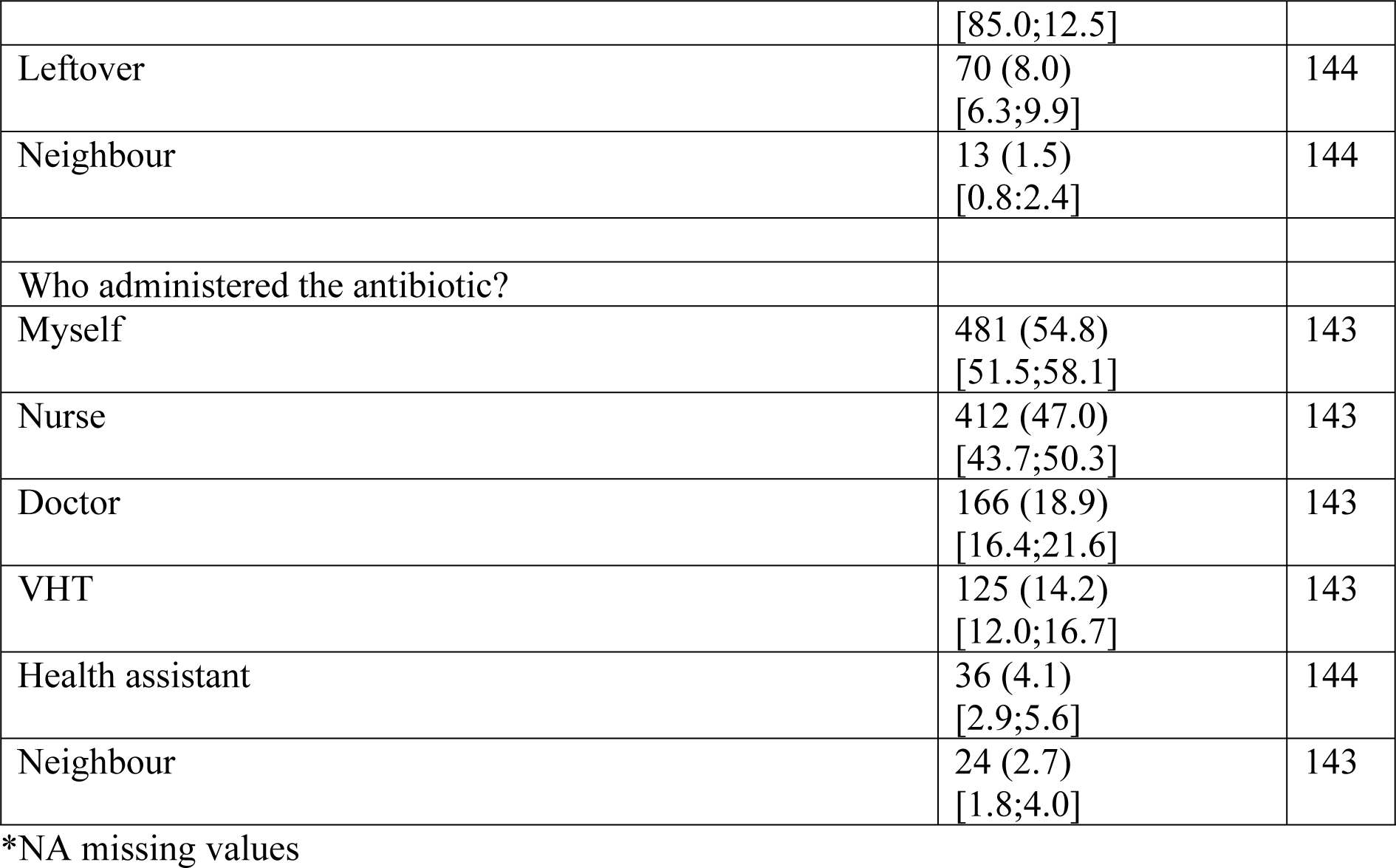
Prescription, source and administration of antibiotics in humans

**Table 2e.** Attitudes towards antibiotic prescription in humans and patient-doctor relationship

| Question | Ministry or farming |  |  | Total | NA* |
| --- | --- | --- | --- | --- | --- |
|  | MAAIF | MOH | Farmer |  |  |
|  | «yes»<br>n (%)<br>[95% CI] | «yes»<br>n (%)<br>[95% CI] | «yes»<br>n (%)<br>[95% CI] | «yes»<br>n (%)<br>[95% CI] |  |
| Did you seek medical help? | 13 (61.9)<br>[40.7;80.0] | 135 (52.7)<br>[46.6;58.8] | 329 (57.5)<br>[53.4;61.5] | 477 (56.1)<br>[52.8;59.4] | 172 |
| I have experienced antibiotic prescription for myself or my kin or my animals/birds. | 19 (61.3)<br>[43.8;76.8] | 227 (75.7)<br>[70.6;80.3] | 469 (75.4)<br>[71.9;78.7] | 716 (75.0)<br>[72.2;77.7] | 68 |
| When antibiotics are prescribed (whether for myself or my animals), the doctors take time to provide information on how they should be used, in an understandable manner. | 22 (84.6)<br>[67.4;94.6] | 252 (84.6)<br>[80.1;88.3] | 520 (82.4)<br>[79.3;85.2] | 795 (83.1)<br>[80.7;85.4] | 66 |
| I usually know how antibiotics should be used/taken, even if I was given information about their use. | 15 (46.9)<br>[30.5;63.8] | 161 (53.5)<br>[47.8;59.1] | 301 (47.7)<br>[43.8;51.6] | 478 (49.5)<br>[46.4;52.7] | 57 |
| I often know before I visit a doctor, whether I need antibiotics or not. | 11 (36.7)<br>[21.3;54.5] | 126 (41.4)<br>[36.0;47.0] | 319 (50.1)<br>[46.2;53.9] | 457 (47.0)<br>[43.9;50.1] | 50 |
| Doctors always conduct a thorough examination regarding whether a patient/animal is in need of antibiotics or not. | 24 (75.0)<br>[58.3;87.4] | 258 (83.8)<br>[79.3;87.6] | 490 (76.8)<br>[73.4;79.9] | 772 (78.8)<br>[76.2;81.3] | 43 |
| Doctors prescribe antibiotics when a patient expects it. | 12 (37.5)<br>[22.4;54.7] | 97 (32.5)<br>[27.4;38.0] | 335 (52.6)<br>[48.7;56.4] | 445 (46.0)<br>[42.8;49.1] | 54 |
| I am confident in a doctor's decision if he/she does not prescribe antibiotics. | 23 (71.9)<br>[54.9;85.1] | 232 (77.1)<br>[72.1;81.5] | 501 (78.5)<br>[75.2;81.6] | 757 (77.9)<br>[75.2;80.4] | 50 |
| A doctor who does not prescribe antibiotics when the patient thinks that they are needed, is not a good doctor. | 7 (22.6)<br>[10.7;39.3] | 85 (28.2)<br>[23.4;33.5] | 371 (58.8)<br>[54.9;62.6] | 464 (48.1)<br>[45.0;51.3] | 58 |
| I usually know how infections should be treated. | 17 (54.8)<br>[37.5;71.3] | 146 (47.7)<br>[42.1;53.3] | 277 (44.0)<br>[40.2;47.3] | 441 (45.6)<br>[42.5;48.7] | 55 |
| I have experience, as patient or a kin, of acquiring prescribed antibiotics from pharmacy. | 20 (64.5)<br>[47.0;79.5] | 211 (70.8)<br>[65.5;75.7] | 434 (69.0)<br>[65.3;72.5] | 665 (69.3)<br>[66.4;72.2] | 63 |
| Pharmacy staff take their time to inform me on how antibiotics should be used. | 16 (64.0)<br>[44.5;80.5] | 152 (55.5)<br>[49.5;61.3] | 371 (68.4)<br>[64.4;72.2] | 540 (64.1)<br>[60.8;67.3] | 180 |

\*NA missing values

Regarding awareness of antibiotic resistance, nearly 80% agreed that antibiotic resistance and fake antibiotics are big problems in Uganda. About 52% are confident that pharmaceutical companies will be able to develop new medicine which will solve the problem. Less than half of the respondents have experienced a sensitization campaign (36.5%) or are aware of organisations involved in minimising the development of antibiotic resistance.

### Knowledge, attitude and practice towards antibiotic access, usage and disposal (questions Q1 to Q9)

The proportions of the correct, wrong and “don’t know” answers are displayed in Table 3. The first three questions Q1 to Q3 asking about acquiring antibiotics without visiting a doctor and confirming the dosage before administration were answered correctly with a clear majority above 70%. Regarding Q4 and Q5 asking about usage and disposal of leftover antibiotics, less than 60% answered correctly. The majority of the respondents (70%) took the wrong answer regarding the questions Q6 that antibiotics fasten recovery for any sickness. More than half of the respondents (52.8%) agreed with the statement in Q7 “as soon as you have fever you should have an antibiotic”. Similarly, in Q8, a slight majority of 55.6% will not “see if the infection goes away on its own”, assuming that antibiotics will be taken immediately after onset of clinical symptoms. Most of the respondents (68.2%) agreed with Q9 that “the body can usually fight mild infections on its own without antibiotics”.

**Table 3.** Manifest variables (questions) to assess the attitude towards antibiotic access, usage and disposal

| Questions | Answers |  |  |
| --- | --- | --- | --- |
|  | Correct | Wrong | Don't Know |
|  | n (%) | n (%) | n (%) |
|  | [95% CI] | [95% CI] | [95% CI] |
| Q1. I think that it is good to be able to acquire antibiotics from relatives or acquaintances, without having to be examined by a doctor.<br>(13 NA*) | 739 (73.2)<br>[70.6;76.0] | 210 (20.8)<br>[18.1;23.6] | 60 (5.9)<br>[3.3;8.7] |
| Q2. I think that it is good to be able to buy antibiotics over the counter, without having to see a doctor. (19 NA*) | 784 (78.2)<br>[75.7;80.7] | 182 (18.1)<br>[15.6;20.7] | 37 (3.7)<br>[1.2;6.2] |
| Q3. I confirm dosage before administration of antibiotics. (15 NA*) | 791 (78.5)<br>[76.1;81.1] | 196 (19.5)<br>[17.0;22.0] | 20 (2.0)<br>[0;4.5] |
| Q4. Leftover antibiotics can be saved for personal future use or to give to someone else. (11 NA*) | 569 (56.3)<br>[53.1;59.5] | 386 (38.2)<br>[35.0;41.4] | 56 (5.5)<br>[2.4;8.8] |
| Q5. I dispose my leftover antibiotics with other household garbage or even in the farm. (15 NA*) | 563 (55.9)<br>[52.7;59.1] | 402 (39.9)<br>[36.7;43.1] | 42 (4.2)<br>[1.0;7.4] |
| Q6. Antibiotics make one recover faster when having any disease. (10 NA*) | 240 (23.7)<br>[20.9;26.6] | 716 (70.7)<br>[68.0;73.6] | 56 (5.5)<br>[2.8;8.4] |
| Q7. As soon as you have fever you should have an antibiotic. (14 NA*) | 451 (44.7)<br>[41.6;48.0] | 532 (52.8)<br>[49.6;56.0] | 25 (2.5)<br>[0;5.8] |
| Q8. If I get an infection, I often wait and see/rest and take it easy, and see if the infection goes away on its own. (23 NA*) | 427 (42.7)<br>[39.5;46.0] | 556 (55.6)<br>[52.4;58.9] | 16 (1.6)<br>[0;4.8] |
| Q9. The body can usually fight mild infections on its own without antibiotics. (14 NA*) | 688 (68.2)<br>[65.4;71.2] | 293 (29.1)<br>[26.2;32.0] | 27 (2.7)<br>[0;5.7] |

\*NA missing values

### Polytomous latent class analysis: questions Q1 to Q9

For the polytomous latent class analyses, the answers of 952 respondents with complete answers were included. Based on BIC and the attempt to have a least a proportion of 0.1 in each class, the best latent class model comprised three classes with the predicted class probabilities of 68.5 % (class I), 13.1 % (class II) and 18.3 % (class III) displayed in Figure 1. The classes were named “accurate knowledge”, “moderate knowledge and restricted use” and “moderate knowledge and relaxed use” because the highest proportion of correct answers for all questions - with the exception of Q8 - were found in class I, the highest proportion of “don’t know” answers were found in class II and the highest proportion of wrong answers was found in class III.

**Figure 1.**
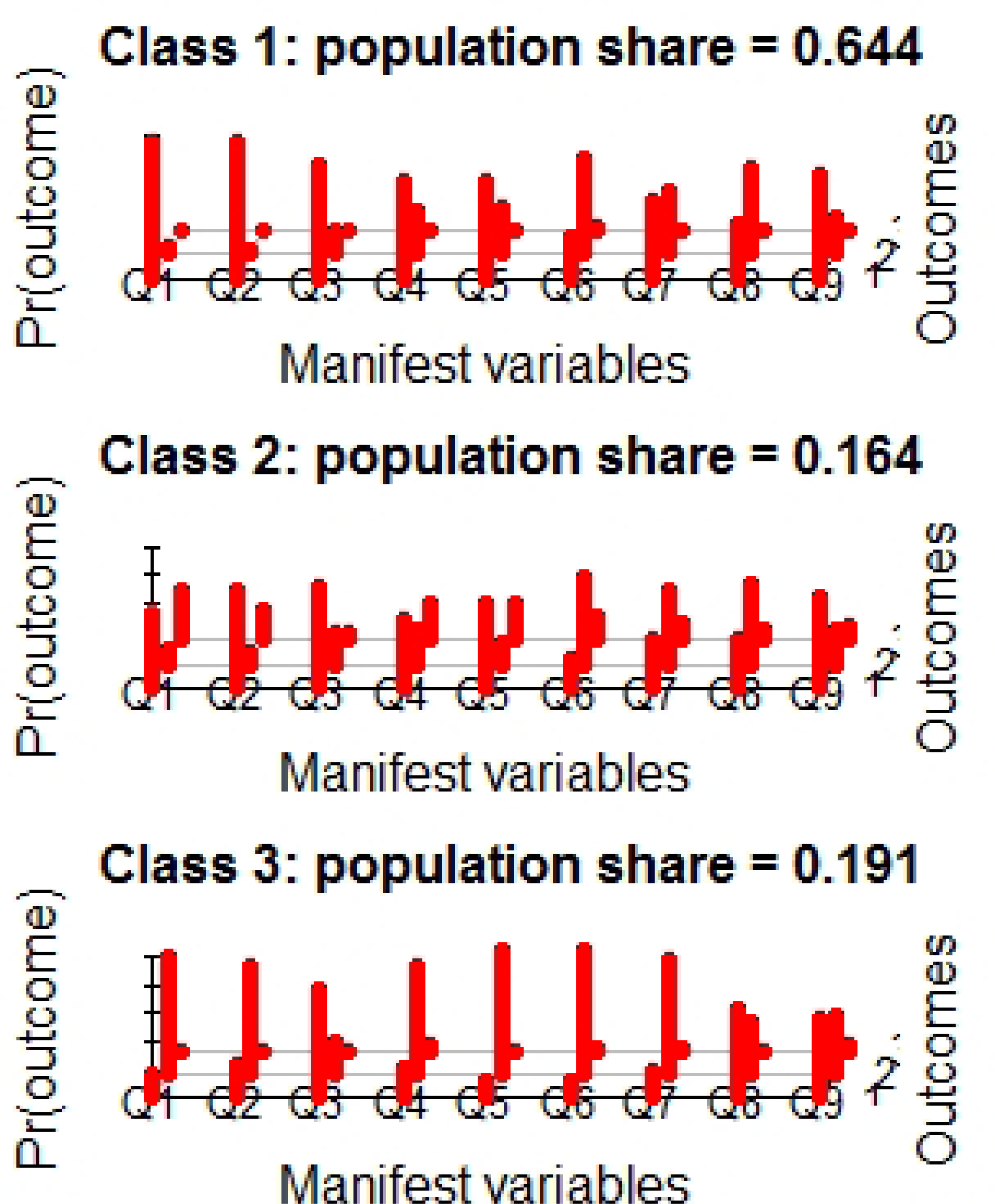
Results of the polytomous latent class analysis for the manifest variables (questions) to assess the attitude towards antibiotic access, usage and disposal

### Multinomial regression for predicted class membership: questions Q1 to Q9

Based on the multinomial regression, assessing potential associations between latent class membership and nine predictors, the following results were obtained (S5 Table). Intensive farmers compared to no farming activity were more likely to be in class II than in class I. Respondents affiliated with MAAIF or MOH compared to respondents not affiliated with these ministries were more likely in class I than in class III. Gender was not significantly associated with class membership. Respondents with increased levels of education compared to primary education were in general more likely to be in class I than in class III or II. Respondents, which keep cattle or goats, were - compared to respondents not keeping these species - more likely to be in class III or class II and III than in class I. Respondents which keep poultry were – compared to those not keeping poultry – more likely in class I than in class II or III. Keeping pigs or sheep was not significantly associated with latent class membership.

Similarly, in a conditional inference tree including all nine potential factors, ministry, education, and keeping cattle were found to be significantly associated with class membership (Figure 2). Respondents affiliated with MAAIF or MOH compared to respondents not affiliated with one of these two ministries were less likely to be on class III. Amongst respondents not affiliated with either MAAIF or MOH, having a higher than a primary education was associated with a higher probability of being in class I. In respondents being not affiliated with MAAIF or MOH and having at least an ordinary education, keeping cattle was associated with a lower proportion of correct and a higher proportion of “don’t know” answers.

**Figure 2.**
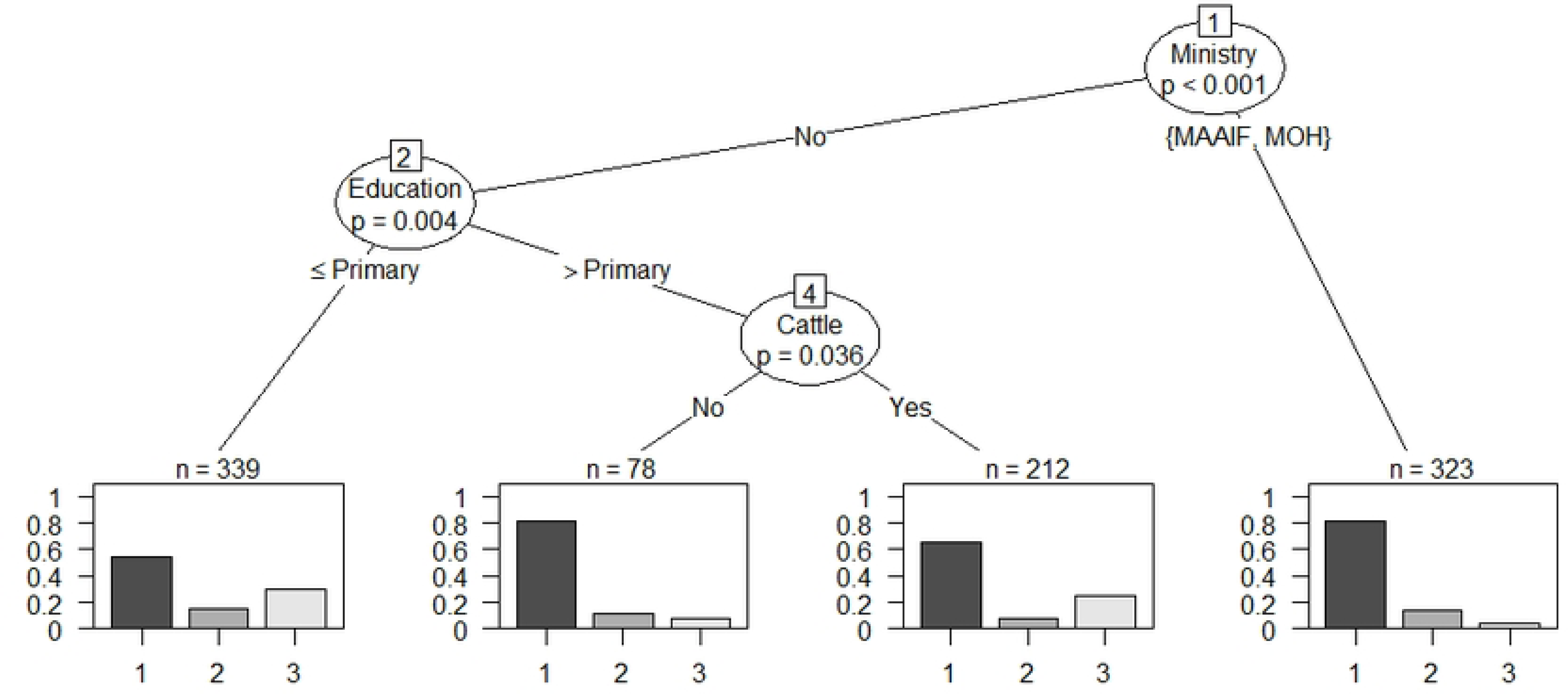
Conditional inference tree for predicted class membership and association with risk fact factors

### Knowledge, attitude and practice towards resistance (questions Q10 to Q17)

The proportions of the correct, wrong and “don’t know” answers are displayed in Table 4. Nearly half of the respondents did not choose the correct answer for questions Q10 and Q11 related to the usage of antibiotics for food preservation or cooking. A majority assumed that antibiotics can prevent sickness in Q12. Correctly answered by a majority was the question Q13 related to usage of animal drugs in humans and vice versa. A majority agreed with the wrong statement in Q14 to take antibiotics any time of sickness. In contrast, a majority took the correct answer in Q15 regarding the question to stop antibiotics when feeling well again. The majority of the respondents agreed with the statement in Q16 to switch the antibiotic if another type has failed. More than 40% of the respondents either assumed that a “cocktail of antibiotics is a solution to resistance” or were uncertain.

**Table 4.** Manifest variables (questions) to assess the attitude towards antibiotic resistance

| Questions | Answers |  |  |
| --- | --- | --- | --- |
|  | Correct | Wrong | Don't Know |
|  | n (%) | n (%) | n (%) |

|  | [95% CI] | [95% CI] | [95% CI] |
| --- | --- | --- | --- |
| Q10. I have heard/learnt that antibiotics can be used to preserve food especially milk. (17 NA*) | 548 (54.5)<br>[51.3;57.8] | 209 (20.8)<br>[17.6;24.1] | 248 (24.7)<br>[21.5;28.0] |
| Q11. I have heard/ learnt that antibiotics quicken and soften food when cooking especially beans and meat. (17 NA*) | 459 (45.7)<br>[42.4;49.0] | 436 (43.4)<br>[40.1;46.7] | 110 (10.9)<br>[7.7;14.3] |
| Q12. Antibiotics can prevent sickness. (11 NA*) | 392 (38.8)<br>[35.7;42.0] | 589 (58.2)<br>[55.2;61.5] | 30 (3.0)<br>[0;6.2] |
| Q13. I think it is okay to use animal drugs in humans and vice versa. (11 NA*) | 765 (75.7)<br>[73.1;78.3] | 157 (15.5)<br>[12.9;18.1] | 89 (8.8)<br>[6.2;11.4] |
| Q14. One can take antibiotics anytime of sickness.(13 NA*) | 393 (38.9)<br>[35.9;42.2] | 588 (58.3)<br>[55.2;61.5] | 28 (2.8)<br>[0;6.0] |
| Q15. I can stop antibiotics anytime I feel well from sickness. (13 NA*) | 629 (62.3)<br>[59.3;65.4] | 341 (33.8)<br>[30.7;36.9] | 39 (3.9)<br>[0.8;7.0] |
| Q16. One can switch to another type of antibiotics if another type has failed.(20 NA*) | 213 (21.2)<br>[18.7;24.0] | 752 (75.0)<br>[72.4;77.8] | 37 (3.7)<br>[1.1;6.4] |
| Q17. Cocktail of different antibiotics is a solution to resistance. (15 NA*) | 573 (56.9)<br>[53.7;60.1] | 239 (23.7)<br>[20.5;26.9] | 195 (19.4)<br>[16.2;22.6] |

\*NA missing values

### Polytomous latent class analysis: questions Q10 to Q17

Based on BIC and the attempt to have at least a proportion of 0.1 in each class, the best latent class model comprised three classes with predicted class probabilities of 57.5 % (class I), 17.7 % (class II) and 24.8 % (class III) displayed in figure 3. These classes were named “fair knowledge”, “moderate” and “low knowledge” as (with the exception of Q16), class I comprised the highest proportion of the correct and the lowest proportion of wrong answers. Class II had the highest proportion of “don’t know” answers and class III had the highest proportion of wrong answers for most of the questions.

**Figure 3.**
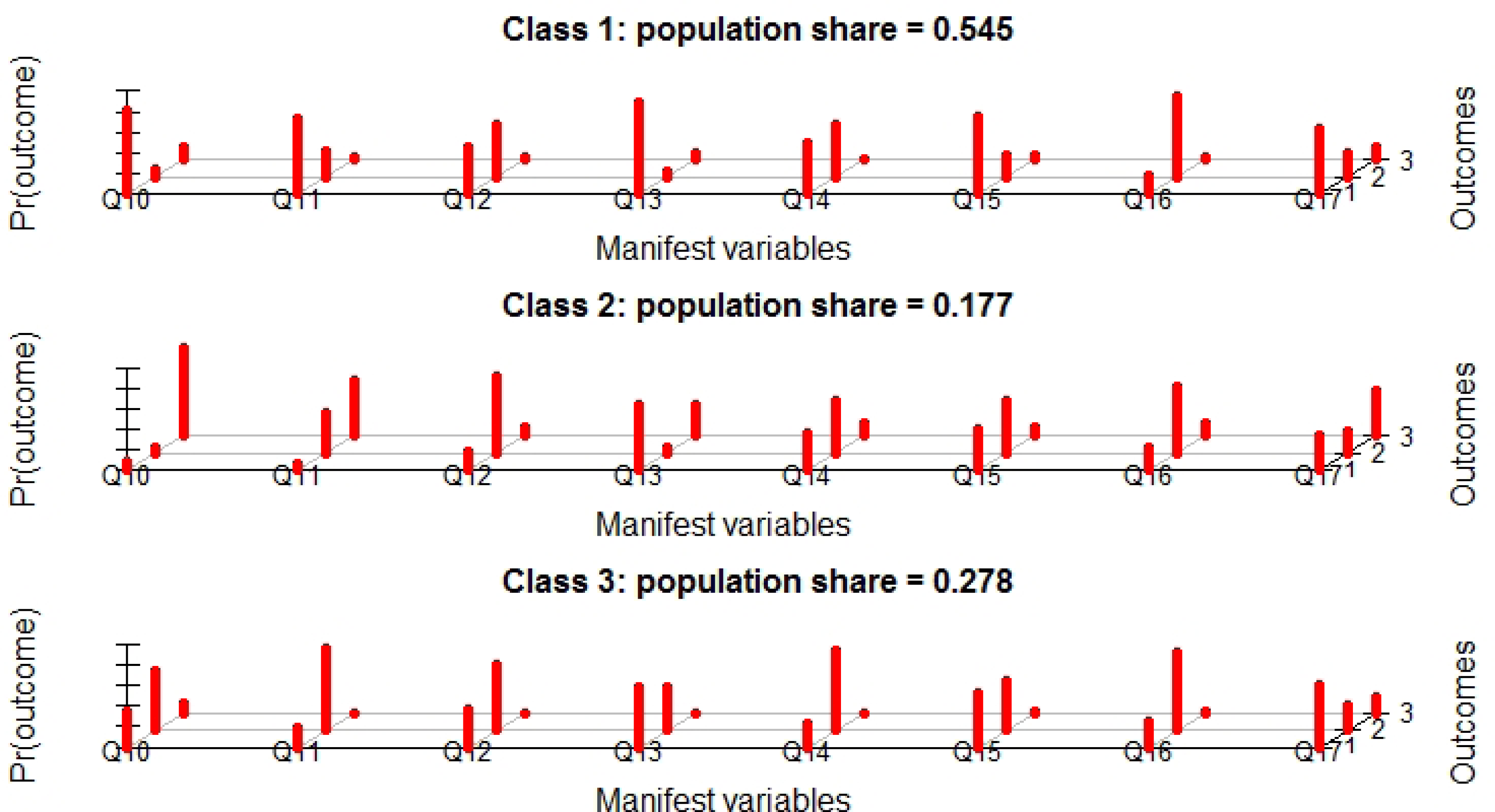
Results of the polytomous latent class analysis for the manifest variables (questions) to assess the attitude towards antibiotic resistance

### Multinomial regression for predicted class membership: questions Q10 to Q17

Based on the multinomial regression assessing potential associations between latent class membership and nine predictors, the following results were obtained (S6 Table). Being a farmer, intensive or semi-intensive, compared to no farmer was not significantly associated with class membership. Respondents affiliated with MAAIF or MOH were less likely to be in class II or III, respectively compared to class I. Females compared to males were more likely in class III than I. Respondents with higher levels of education (tertiary), compared to primary education, were more likely to be in class I, than in class III. Keeping cattle, compared to not keeping cattle, was associated with being more likely in class II than class I. Keeping pigs compared to not keeping pigs, was associated with being more likely in class I compared to II. Keeping poultry, compared to not keeping poultry, was associated with being more likely in class III than in class I. Respondents keeping sheep were more likely in class 1 than in classes II and III.

Based on a conditional inference tree, keeping poultry, sheep cattle or goats as well as type of farming and education level was found to be significantly associated with predicted class membership (Figure 4). Keeping poultry was associated with a lower probability of being in class I, unless respondents had tertiary or university education. Amongst poultry keepers with primary or ordinary education, keeping goats was associated with a higher proportion of class I members. Amongst those who did not keep poultry, being a semi-intensive farmer compared to intensive or no farming was significantly associated with class membership. Amongst semi-intensive farmers keeping sheep and cattle was significantly associated with class membership. While keeping sheep was associated with a higher probability of class I, keeping cattle was associated with a lower probability of class I membership.

**Figure 4.**
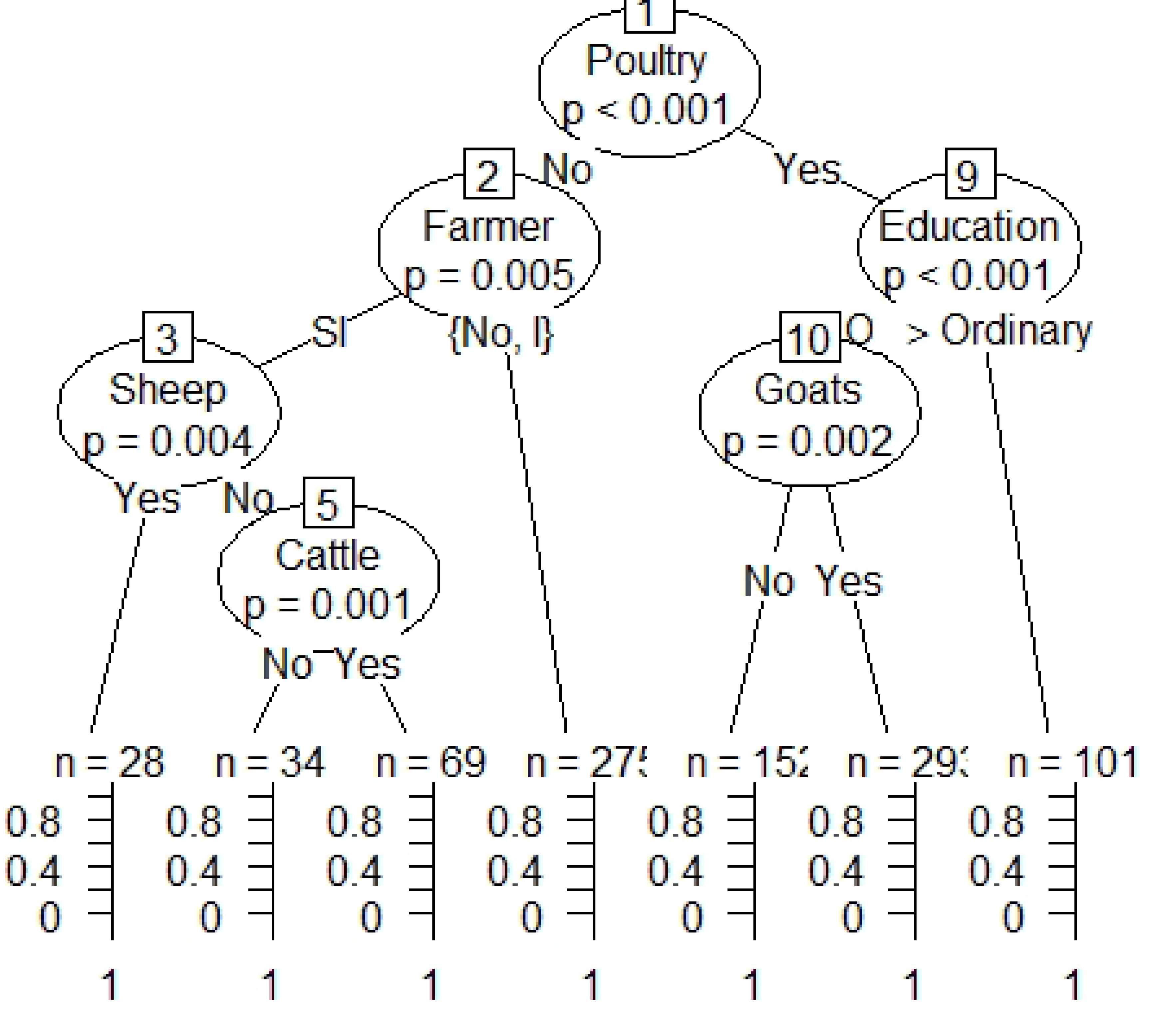
Conditional inference tree for predicted class membership and association with risk fact factors

## Discussion

This study assessed knowledge, attitudes and practices related to antibiotic resistance, access and usage in both animals and humans amongst respondents being affiliated with Uganda’s Ministry of Health (MOH) and Ministry of Agriculture, Animal Industry and Fisheries (MAAIF) or those with a background of farming in Kyegegwa, Uganda. This is one of the few studies undertaken in LMICS, particularly in East Africa. The respondents of this are key players in the prescription and use of antimicrobials in humans and livestock. Misuse of antimicrobials is reported as critical in the development and spread of antimicrobial resistance (22,23) and this can be perpetuated by uninformed users or reckless professionals.

In this study amoxicillin, penicillin, erythromycin and co-trimoxazole were found to be the most commonly used antibiotics in humans. This is similar to what is reported in other studies in Tanzania (24,25), Ethiopia (26), Uganda (27,28), Sudan (29). However, some studies report metronidazole as the most commonly used antibiotic (30). A number of factors in LMICS influence the decision to self-medicate. These include poverty, long distance to health facilities, easy access to drugs from pharmacies and drug shops, mild illness, ignorance, poor attitudes of health workers, knowledge of diagnosis (retreatment of similar illness), convenience, and non-availability of doctors (28,31,32).

In 39.1% members of the respondents’ households of this study were on antibiotics, a manifestation of the large-scale use of antibiotic in Uganda. Self-medication is so widespread that WHO now considers it as a component of self-care (32). Self-medication is one of the major drivers of drug resistance (33,34) and reflects a lack of access to appropriate health care. The commonly used drugs in self-medication are prone to increased resistance, especially when inappropriately used.

For the livestock sector, penicillins, tetracyclines and gentamycin were mentioned as the most commonly used. Some studies in Africa and elsewhere have reported similar results and trends (35–37). Unfortunately, similar studies in LMIC are still scarce, particularly those focussing on prescribers. Several factors drive farmers and livestock keepers to self-medicate their animals. This can be attributed to readily available, easy access, cheaper drugs, weak regulatory measure and lack of qualified personnel. One of the main drivers to use antibiotics is commercial both by the suppliers and the consumers. Livestock farmers and animal keepers want their animals healthy and productive.

Since just a subset of all respondents kept animals in this study, comparing the frequencies in humans and animals is only of limited sense. However, it is worthwhile mentioning, that all antibiotics used in humans are also used in animals. Surprisingly high was the frequency of antibiotics given to cattle, even higher than to any human age category. This might be explained by part of the study region lying in the cattle corridor, a zone stretching from south-west to north-east Uganda, dominated by pastoral rangelands, and cattle being very valuable compared to other species.

These data are self-reported frequencies and need therefore to be carefully interpreted in terms of a potential social desirability bias. What is yet missing in Uganda is a national surveillance system to monitor antibiotic consumption in animals and humans. Some efforts have been put in place by Ministries of Health and Agriculture and a National Antimicrobial Resistant Action plan for Uganda has been drafted. These action plans have been developed within the framework of tripartite agreements by the Food and Agriculture Organization (FAO), World Health Organization (WHO) and World Organization for Animal Health (WOAH) (38,39). It is desirable to understand the amount of antibiotics used both in livestock and humans as well as their effects on humans, animals and the environment, and the drivers of resistance in order to improve on the rational use.

In this study, the majority of respondents in all categories have experienced prescription and contented, that doctors provide adequate advice. The fact, that a majority seek prescription from a doctor is a positive attribute and a good prospect of judicious antibiotic use, however these results need to be interpreted with caution, since in many communities in Uganda, the word ‘doctor’ is more or less synonymous with health worker or provider, regardless of the level of qualification and/or position in the health care service provisions.

Alarming is the high proportion of 82.9% of the respondents who indicated that they were not following the recommendation of withholding periods when administering antibiotics to livestock. Thus, milk and meat containing antibiotic residues from treated animals potentially enter the food chain and pose risks to consumers. Similar findings of not observing the withholding period were also observed in animals in Tanzania (40). Presumably, the lack of observing a withholding period is not due to a lack of knowledge, but due to financial constraints and a lack of control for residues along the food chain (41). Antibiotics will continue to be used in livestock production, particularly to treat and control pathogenic infections. But lack of observance of withholding periods will potentially pose public health hazards in terms of effects of drug residues like carcinogenicity and toxicity associated with consumption of animal products and development of antimicrobial resistance.

About a third (32.1%) of the respondents indicated administering antibiotics for growth and fattening. This is slightly higher than what is reported in a neighbouring country of Uganda, Rwanda, 26.5% (42). Currently, in Uganda, officially there is a ban on the use of antibiotics for growth promotion in livestock. In Africa, a recent survey by the World Organisation for Animal Health (43) found out that 15% of countries who participated, still use antibiotics for growth promotion. Use of antibiotics for growth promotion normally entails use of lower concentrations of the active ingredients over a longer period and this puts an extensive pressure leading to emergence of antimicrobial resistance strains of bacteria (44). Consequently, a number of countries have banned use of antibiotics for growth promotion.

Following the approach from (16) we assessed knowledge by analysing response patterns to a number of questions with a clustering approach. A first set of nine questions was dedicated to assess knowledge and attitude towards antibiotic access, usage and disposal. Based on a polytomous latent class analysis we obtained three classes which we named “class I: accurate knowledge” (68.5%), “class II: moderate knowledge & restricted use” (13.1%) and “class III moderate knowledge and relaxed use”. A clear majority of respondents in class I answered correctly questions related to unrestricted antibiotic access and dosage confirmation, thus giving evidence that unrestricted access is perceived as a driving factor for antibiotic resistance. Less respondents answered correctly questions related to saving leftover antibiotics for future use. This might be contextual and be explained by potential anticipated antibiotic shortage. Similarly, the questions related to disposal of leftover with the household garbage or even on the farm might indicate the absence of appropriate waste management. Worrisome are some misconceptions, also being present in the class I with the most accurate knowledge with regard to using antibiotics immediately with the onset of fever and the assumption that recovery for any disease is achieved earlier with antibiotics.

Based on the discussions during the dialogue days, a symposium in the context of a collaboration between Makerere university and university of Zurich, it became evident that not all questions from (16) are directly applicable to the situation in Uganda. Participants mentioned (mis-)use of antibiotics to preserve milk in the absence of suitable storage conditions, to obtain “stronger” self-brewed alcoholic beverages or to soften meat and beans when cooking. These practices are particularly worrisome and should be considered in sensitization campaigns.

## Conclusions

Inadequate knowledge, inappropriate attitudes and practices along the antimicrobial supply is a potential danger to the development and spread of antimicrobial resistance. Similar generic drugs or surrogates are used both in human and animal treatment, moreover with unrestricted access and self-prescriptions if not appropriately addressed will exacerbate the challenges of antimicrobial resistance. One of the weakest points in the use of antimicrobial is enforcement of the existing laws and guidelines. This can be complicated by poverty, ignorance and other socio-economic factors. Evidence generated in this study should support further policy formulations and strategies to regulate AMU and implementation of local and national action plans. It can be used in sensitizations and promotions of judicial use of antibiotics in livestock and communities, encouraging alternative options for disease control in livestock like promotion of vaccinations. A key component of combating the increase of antibiotic resistance will require a shift and promotion of One Health approach, that promotes intersectoral, multidisciplinary collaborations and effective communication.

## Data Availability

All relevant data are within the manuscript and its Supporting Information files.

## Acknowledgements

To the respondents.

Special appreciation goes to Kyegegwa district extension staff with which data for this report thesis was successfully collected. Field supervisor Mr. Friday Kyomya.

## Author contributions

Conceived the study: MAK, SH, MK, DO, CK, TO. Designed the questionnaire: MAK, DO, TO, SH. Tested and applied the questionnaire: MAK. Analyzed the data: MAK, SH. Wrote the paper: SH, TO, MAK, MK, DO.

## Supporting information

S1 Questionnaire in English

S2 Data

S3 Code for analysis

S4 Table Results of the multinomial logistic regression to assess the association between demographic factors and latent class membership of attitude towards antibiotic access, usage and disposal

S5 Table Results of the multinomial logistic regression to assess the association between demographic factors and latent class membership of attitude towards antibiotic resistance

